# Pretransplant BK virus-specific T-cell-mediated immunity and serotype specific antibodies may have utility in identifying patients at risk for BK virus associated haemorrhagic cystitis after allogeneic HSCT

**DOI:** 10.1101/2021.06.09.21258555

**Authors:** Markéta Šťastná-Marková, Eva Hamšíková, Petr Hainz, Petr Hubáček, Marie Kroutilová, Jitka Kryštofová, Viera Ludvíková, Jan Musil, Pavla Pecherková, Martina Saláková, Vojtěch Šroller, Jan Vydra, Šárka Němečková

## Abstract

BK polyomavirus (BKV) persists lifelong in the urinary tract with asymptomatic urinary shedding in healthy individuals. In immunocompromised persons after transplantation of hematopoietic stem cells (HSCT) the BKV high-rate replication is associated with haemorrhagic cystitis (HC) with a reported incidence of 17 %. Numerous studies of reconstitution of the immune system after HSCT have established the principal role of T cell effectors in the control of viral replication and reactivation. The value of pretransplant BKV-specific antibodies in transplanted patients for the protection from viral disease was long considered insignificant. We hypothesized that the status of BKV immunity prior to HSCT could provide evidence for the BKV tendency to reactivate and that examining the level of subtype-specific antibodies and T-cell response in individual patients could help to predict the risk of BKV reactivation and HC. Evaluation of the risk of HC in relation to pretransplant anti-BKV1,2,4 IgG levels together with clinical factors known before transplantation revealed that patients with „medium” anti-BKV IgG and significant clinical risk (SR) have a very significantly increased HC risk in comparison with the reference group of “low” anti-BKV IgG level and low clinical risk (LR) (P=0.0009). Predictive value of pretransplant BKV specific IgG was confirmed on the level of virus genotypes. Analysis of pretransplant T cell immunity to BKV antigens VP1 and LTag has shown that magnitude of IFN-gamma T cell response inversely corelated with posttransplant DNAuria. We hypothesize that the control of BKV latency by BKV specific T cells before HSCT would be one of the factors that influence BKV reactivation after HSCT. Our study has shown that prediction using a combination of clinical and immunological pretransplant risk factors can help early identification of patients who are at risk of developing BKV disease after HSCT.

## INTRODUCTION

Human BK polyomavirus (BKV) is a highly prevalent virus that establishes latency in the urinary tract after usually asymptomatic primary infection early in life. BKV isolates are classified into four major subtypes based on the VP1-DNA sequence. Subtype I is prevalent worldwide (80%), followed by subtype IV (15%), which is common in Asia and parts of Europe. Subtypes II and III occur less frequently [1]. Seroepidemiology studies have shown that antibody responses specific for subtype I, including all subgroup variants, can be found in up to 98% of adults [2]. Seroreactivity to subtypes II, III and IV in healthy donors was 86%, 77% and 80%, respectively. Detection of high seroprevalence of rare serotypes II and III could be ascribed to cross-reactivity with serotype IV. Cross-reactivity was also observed among subgroups of serotype I [3]. A seroepidemiologic study of BKV in the Czech Republic has shown that the seropositivity rate of 80% found for young adults decreased with age to 56% [4], indicating that virus latency can be associated with the waning of specific humoral immunity. BKV infection is not associated with any pathology in immunocompetent individuals, but in immunocompromised patients after transplantation of kidney or of hematopoietic stem cells (HSCT), BKV reactivation/reinfection can cause serious clinical illness. BKV reactivation after allogeneic HSCT can cause haemorrhagic cystitis (HC) or nephropathy (For an overview, see [5]. BKV infection is responsible for haemorrhagic cystitis (HC) in up to 17% of recipients of allogeneic HSCT within the first year after treatment [6]. BK virus DNA can be detected in the urine of up to 80% of patients after allogeneic HSCT [7, 8]. However, patients who develop symptoms of HC excrete significantly higher levels of BKV DNA in urine [9]. Reported risk factors of HC are associated with immunosuppression and damage to the urinary tract epithelium by chemotherapy. They include cord blood HSCT, myeloablative conditioning (MAC), chemotherapy with cyclophosphamide, anti-thymocyte globulin (ATG) administration and graft versus host disease (GvHD) therapy. As no effective antiviral treatment of BKV infection in HSCT patients is currently available, BKV-specific adoptive T cell therapies were developed [10].

The value of pretransplant BKV-specific antibodies in transplanted patients for the protection from viral disease was long considered insignificant. Surprisingly, subtype-specific antibodies against BKV VP1 have been reported to have a subtype-specific virus-neutralizing activity in kidney transplanted patients [11]. The predictive value of pretransplant anti-BKV IgG in allo-HSCT recipients is less evident. Positive pretransplant anti-BKV IgG has been regarded as a significant predictive value for viruria [12-14]. However, it was observed that very high levels of anti-BKV IgG (titer>1:40 960) prior to allo-HSCT can be associated with lower peak viremia in the first 100 days after HSCT, whereas anti-BKV IgG titers below 1:40 960 were associated with higher grade viremia [15].

Cellular immune responses against BKV antigens in HSCT patients were examined in a small number of studies [16, 17] usually after transplantation in context with BKV reactivation and HC. We hypothesized that the status of BKV immunity prior to HSCT could provide evidence for the BKV tendency to reactivate and that examining the level of subtype-specific antibodies and T-cell response in individual patients could help predict the risk of BKV reactivation and HC. To evaluate the risk of HC in relation to clinical factors known before transplantation, we analysed a large cohort of HSCT recipients. The predictive value of BKV specific immunity in combination with clinical risk factors for HC was then confirmed in a smaller cohort of transplanted patients.

## MATERIAL AND METHODS

### Patients, donors and sample collection in PBIHC study (<u>P</u>retransplant <u>B</u>KV-specific <u>I</u>mmune response for <u>HC</u> Risk assessment)

HSCT recipients (n=149) and their HSC donors (n=120) were invited to participate in the PBIHC study at the Institute of Haematology and Blood Transfusion (IHBT) in the period of 2017-2020. Pretransplant serum/plasma sample of every patient or donor was isolated and stored in frozen state. The patients signed an informed consent form. The study was approved by the institutional ethical board. All transplanted patients obtained peripheral blood progenitor cells as a graft. Urine samples were collected from most of the patients during the first month after HSCT; afterwards, sampling was based on clinical indication. During the PBIHC study, 37 patients donated additional blood sample one week before transplantation for a detection of BKV-specific T cell response.

#### Haemorrhagic cystitis diagnosis

Twenty-two of the 149 patients developed HC grade 2 or higher based on clinical symptoms. Grade 2 HC was diagnosed in three, grade 3 in sixteen and grade 4 in three of them. Additional 12 of the 149 patients had HC grade 1. Grade 1 was defined as microscopic haematuria, grade 2 as macroscopic haematuria, grade 3 as macroscopic haematuria with small clots and grade 4 as gross haematuria with clots and nephropathy [6]. Ninety percent of patients were examined for BKV DNAuria at least once. The BKV viral load was measured by a real-time polymerase chain reaction (qPCR) in the Laboratory of Virology of the Motol Hospital, Prague. The viral load was expressed as copies per ml/urine [18, 19]

#### Assesment of HC risk upon patient demographic and clinical pretransplant characteristics

To identify potential risk factors associated with HC and to determine the predictive value of data concerning age, gender, diagnosis, donor type, conditioning regimen intensity, GVHD, ATG or posttransplant cyclophosphamide (PTCP), we retrospectively analysed clinical data of all 524 patients who underwent allogeneic HSCT at the IHBT in years 2014-2020. The resulting algorithm was afterwards used for prediction of low clinical risk (LR) and significant clinical risk (SR) patients in the PBIHC study. The data of retrospective patient cluster (n=524) are summarized in Table S1 and the analysis is in detail described in supplementary methods.

#### BKV virus-like particles

Production of virus-like particles (VLP) of human polyomaviruses was described previously [20]. Detailed description is in the Ssupplementary Methods.

#### Measurement of BKV specific antibodies

The presence of antibodies to BKV1, 2 and 4 was determined using an in-house enzyme-linked immunosorbent assay (ELISA) based on VP1 virus-like particles (VLP) as described earlier [4, 21]. Pretransplant anti-BKV IgG levels of HSCT recipients were divided into “low”, “medium” and “high”. Detailed description is in the Supplementary Methods.

#### Detection of BKV-specific T cells

PBMCs isolated from the blood of patients and HSC donors were stimulated with peptide pools (PepMix™) covering BKV VP1 and BKV LTag and cultivated in the presence of IL-4 and IL-7. Frequency of the BKV-specific T cells among expanded T cells was measured by ELISPOT-IFNγ or by flow cytometry with intracellular cytokine staining (IC FACS). Detailed description is in the Supplementary Methods.

#### BKV genotyping

Viral subtype identification was performed by sequencing of a region of the VP1 gene as described by Morel V. et al [22]. Detailed description is in the Supplementary Methods. ***Statistics***: Differences in categorical and continuous variables were assessed with the Fisher exact and Mann-Whitney or Wilcoxon matched-pairs signed rank test, respectively. Association of continuous variables was evaluated by the Spearman correlation analysis. Multivariate analysis was performed by using logistic regression model. Cumulative incidence curves were compared using LogRank (Mantel-Cox) test. The tests were conducted at a level of significance higher than 0.05. All calculations and data plots were performed using GraphPad Prism software, version 9.1.0 for Windows, GraphPad Software, USA.

## RESULTS

### Polyomavirus specific antibodies before HSCT in recipients and donors

To find an independent risk marker of the BKV infection and HC, antibodies against BKV1,2,4 and JCV were measured in patients and donors before HSCT (PBIHC study). The specific seropositivity against BKV1, BKV2, BKV4 and JCV was found in 65%, 46%, 41% and 50% of patients and in 65%, 49%, 44% and 47% of HSC donors, respectively. The differences between the medians of the OD index (see Material and Methods) were significant between BKV1 vs. BKV2 or BKV4 (p<0.0001) for both the recipient and donor (Fig. S1 A). Negativity against any BKV found in patients (26%) and donors (28%) was not significantly different (Fisher’s test P=0.6797). No antibodies against any BKV subtype were found in 26% of patients, however, the detection of IFNγ T cell response before HSCT or detection of BKV DNA in urine shortly after HSCT gave evidence that some of those patients had experienced BKV infection. Their seronegativity could most probably be ascribed to a waning specific humoral response, though actual infection cannot be ruled out either. We analysed whether anti-BKV IgG levels were age dependent by correlation analysis (Fig. S1B). The difference between younger (21-45 years) (n=58) and older (46-69 years) (n=58) patients was not statistically significant, with P=0.9636 and P=0.9837 for BKV1 and BKV4, respectively.

We tested whether the level of pretransplant polyomavirus specific antibodies is associated with DNAuria during the first year after HSCT. The maximum level of virus DNA in urine and OD index of virus specific antibodies correlated significantly (Fig. S1B) for BKV4 (Spearman correlation Rs= 0.21; P=0.008) and BKV2 (Rs=0.19; P=0.015). Correlation was not found for BKV1, JCV and donor antibodies of any specificity. The values of antibodies specific for any BKV genotype mutually correlated significantly (P<0.00001). Very high correlation between BKV2 and BKV4 specific antibodies was found in recipient (Rs 0.85) and donor (Rs=0.9) sera. It was most probably caused by high amino acid VP1 sequence homology, and for this reason, the effect of anti BKV2 was not further studied. The role of donor anti-BKV IgG seroprevalence in BKV reactivation was also determined in a group of recipients who were seropositive for BKV1,4 (n=90). Viruria of >10^7^ copies/ml was detected in recipients of grafts from donors with an anti-BKV IgG OD index of <1 or >1 in 37% and 36% of cases, respectively. It implies that donor’
ss anti-BKV immunity does not corelate with the incidence of viruria in HSCT recipient. To confirm a predictive value of pretransplant BKV-IgG, we determined BKV genotypes in samples of 34 HSCT recipients with viruria >10^9^/ml. Sixty-two percent (21) of patients were positive for BKV I/b-2, 11.7% (4) for BKV I/b-1, 2.9% (1) for BKV III and 23.5% (8) for BKV IV/c-2. We observed that higher pretransplant serum antibody levels (BKV1 or BKV4) were directed against that virus genotype which was detected after the HSCT in patient’s urine (Fig. 1C). The differences between serotype specific antibodies were significantly different for patients with BKV I/b-2 and IV/c-2

**Figure 1.**
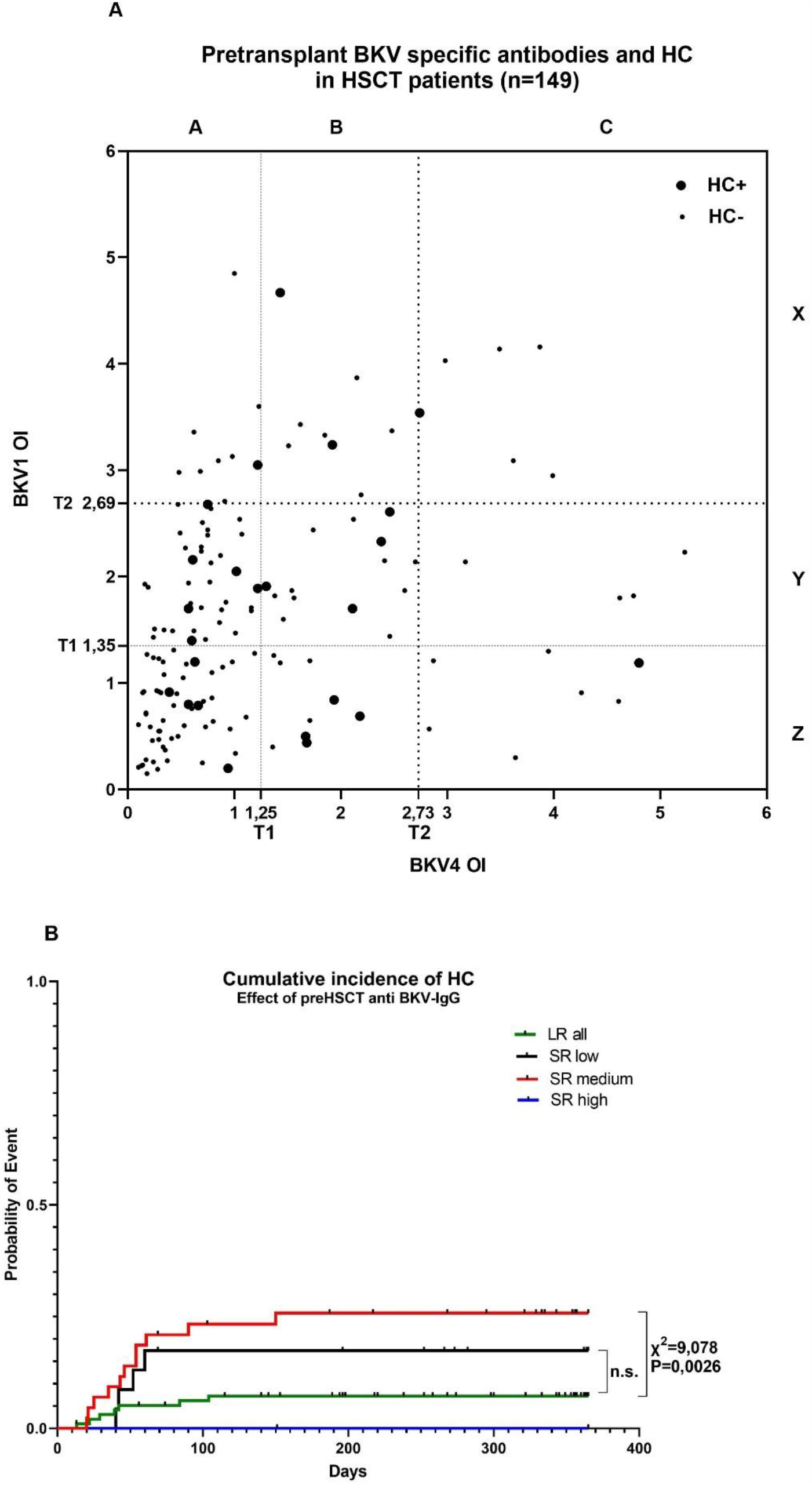
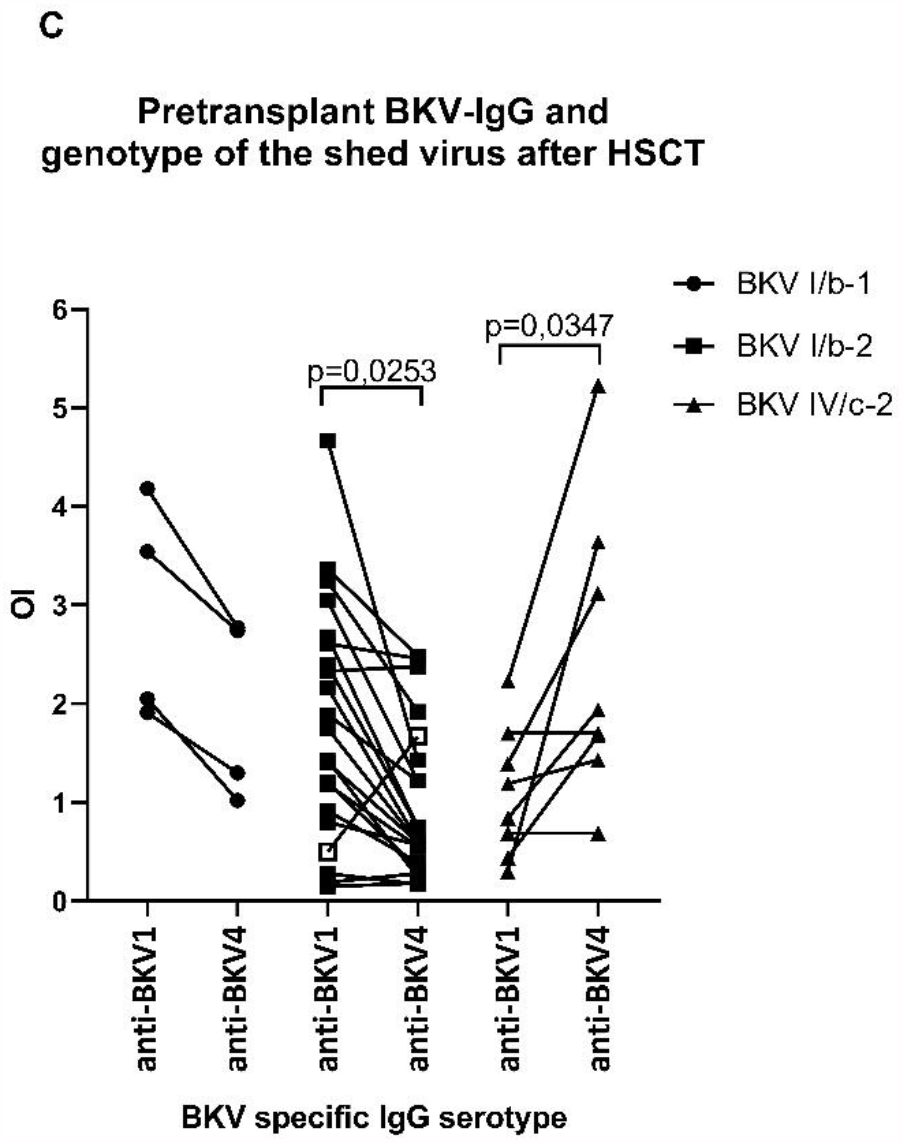
**(A). Plot of pretransplant anti-BKV1,2,4 IgG in individual patients of PBIHC study group (n=149).** Large symbols stand for patients with HC. Dotted line starting at T1 represents the guaranteed threshold of positivity for BKV1 and 4 (mean of nonreactive samples of values < OI=1 + 3 s.d.). Dotted line starting at T2 represents 75% quartile of all seropositive samples > OI=1. **(B) Cumulative incidence of hemorrhagic cystitis (HC) in HSCT recipients stratified according to anti-BKV-IgG levels and clinical risk factors (n=149)**. HC rate in SR group with medium levels of anti-BKV IgG was 34,3%. Survival curves were compared using LogRank (Mantel-Cox) test. **(C) Pretransplant serotype specific IgG correlate with the genotype of excreted virus after HSCT**. The genotype was determined in urine samples of patients with very high BKV load (n=33). Significance of the difference between anti-BKV1 and anti BKV4-IgG levels was determined by the Wilcoxon matched-pairs signed rank test. Reverse ratio between anti-BKV4 and BKV1 IgG observed in one patient (open square) can be possibly ascribed to de novo BKV1 infection that has been detected 7 days before HSCT.

### “Medium” levels of pretransplant BKV specific IgG of patients at significant clinical risk (SR) are associated with increased risk of HC

Further, we asked whether the levels of pretransplant anti-BKV specific IgG of patients affected the incidence of HC. The levels of pretransplant BKV1 and 4 specific IgG antibodies of individual patients are shown in Fig.1 A. Large symbols stand for patients with HC. The overall results (Table 1) show HC to be less common (9.0%) in patients with “low” levels of pretransplant antibodies. This cluster includes seronegative patients with an OD index of <1 as well as those with antibody levels below the T1 threshold level for both BKV1 and BKV4. The group of patients with the highest frequency of HC (19.7%) had “medium” antibody levels. The patients with “high” levels of pretransplant anti-BKV1,4 IgG antibodies were not at increased risk for HC (13.0%). Hence, incidence of HC did not significantly differ between groups. (Table 1). In our study, the “medium” antibody level against any BKV or JCV of donors was not associated with increased risk for HC (Fisher’s exact test P=0,7978) (not shown). To reveal the combined risk for HC, the method for HC risk assessment based upon clinical characteristics verified on a large patient cohort (Table S1) (n=524) was applied to patients from a recent PBIHC clinical study (Table 1) (n=149). For the PBIHC cohort, the difference between SR and LR groups was statistically significant (Table 1, HR=3.362; CI95% = 1.500 to 7.613; P=0.038) in univariate analysis. For overall risk assessment, we considered combined risk emerging from stratification of patients according to pretransplant clinical data (Table S1 and Table 1) and anti-BKV IgG levels. This approach improved HC risk prediction, with the SR „medium” anti-BKV IgG cohort being at a very significantly higher HC risk than the reference LR “low” anti-BKV IgG cohort (Table 1, HR=0.4165; CI95% = 0.2832 to 0.6466; (P=0.0009). Analysis of cumulative incidence of HC in HSCT recipients during the first year after transplantation stratified according to anti-BKV-IgG levels and clinical risk factors revealed significantly higher HC incidence in SR group with medium levels of anti-BKV IgG in comparison with LR group (Fig.1B) (χ^2^=9.078, P=0.0026).

**Table 1.**
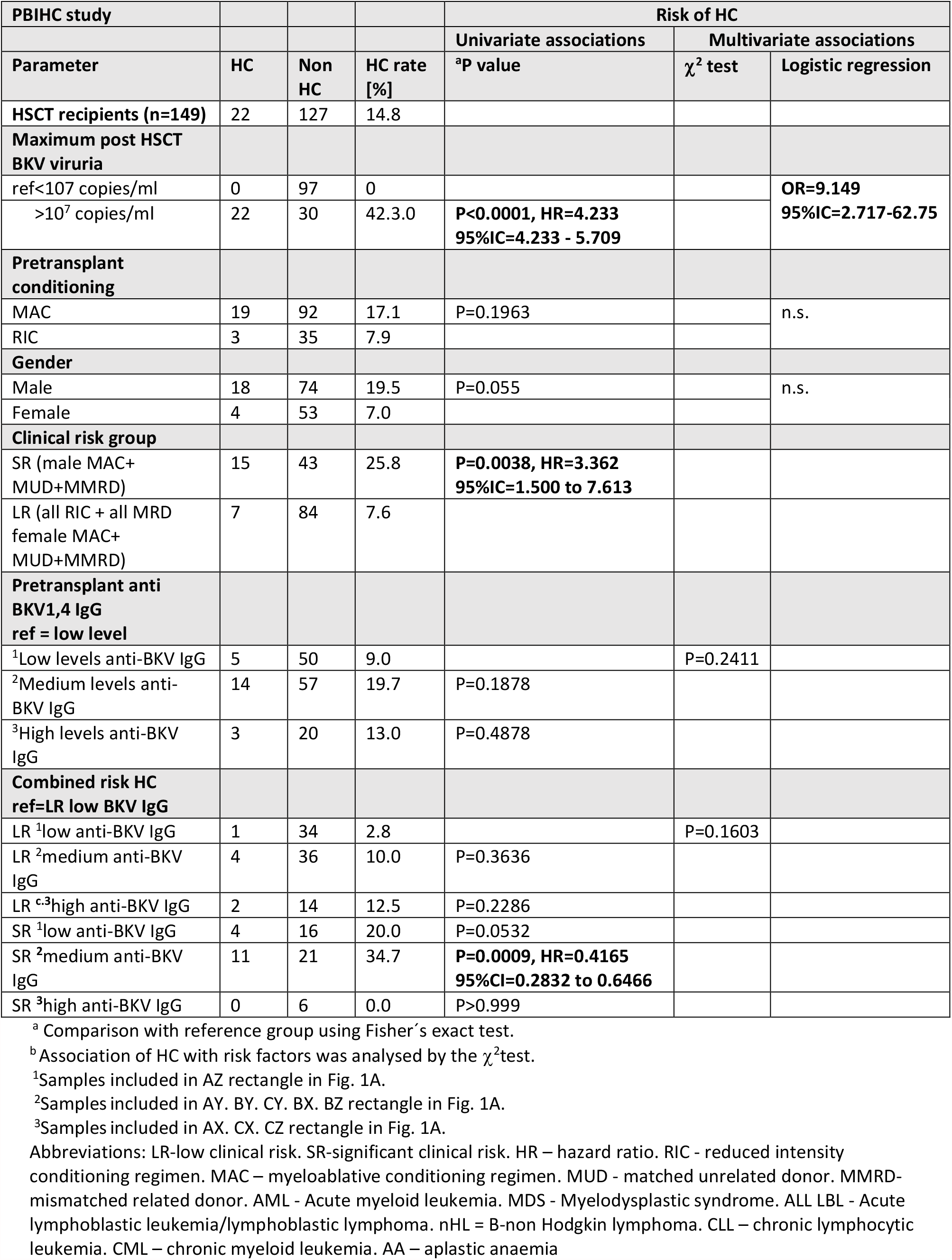
Risk of HC in PBIHC study cohort of HSCT recipients 2017-2020 (n=149) including clinical features and pretransplant BKV specific antibodies

### Pretransplant non-specific and BKV specific T cell response is low in patients with high BKV viruria after HSCT

We wanted to know whether the state of BKV specific adaptive cellular immunity prior to HSCT influenced reactivation of BKV infection. Therefore, in addition to humoral response detection, the T cell response against BKV VP1 and large T antigen was measured in a subgroup of patients (n=34) randomly recruited in the PBIHC study. The evaluation was performed only in the patients with at least one positive anamnestic marker of BKV infection (i.e., in those positive for pre-transplant anti-BKV IgG, early DNAuria or positive pre-transplant T cell response). Before the start of conditioning, the patients were examined for T cell response against VP1 and LTag (Fig. 2A). T cell response was compared between groups of patients stratified according to the posttransplant level of BKV DNAuria. The analysis revealed that T cells of patients with posttransplant DNAuria of <10^7^ BKV DNA copies/ml responded significantly better to stimulation with anti-CD3 (P=0.0111) and with VP1 peptide pool (P=0.0362) than T cells of patients with posttransplant DNAuria of >10^7^ BKV-DNA copies/ml. We hypothesize that the control of BKV latency by BKV specific T cell response before HSCT would be one of the factors that influence BKV reactivation after HSCT. The results of ELISPOT-IFNγ were confirmed by IC FACS-IFNγ in a subgroup of patients with sufficient numbers of *in vitro* expanded cells (n=32). The analysis revealed that BKV VP1 or LTag antigens were recognized by their CD4^+^ and CD8^+^ T cells with significantly prevailing CD4^+^ response (Fig. 2B). Inhibitory molecules PD1 and TIGIT were determined on IFNγ producing T cells responding to stimulation with BKV antigens or anti-CD3 antibody (Fig.2C). The expression of TIGIT (marker of anergy) on BKV specific IFNγ^+^ CD4^+^ T cells restimulated with anti-CD3 or VP1 was significantly decreased if they were from patients with low posttransplant DNAuria (<10^7^ BKV-DNA copies/ml) in comparison with cells of patients with high posttransplant DNAuria of >10^7^ BKV-DNA copies/ml. It could mean that less functional T cell response before HSCT leads to higher viruria after HSCT. Phenotype analysis of T cells also revealed that responding IFNγ^+^ cells were mostly effector memory T cells of phenotype CD45RA^-^RO^+^CCR7^-^ in 80% and 45% of CD4^+^ and CD8^+^ T cells, respectively. Between 15% and 20% were central memory T cells CD45RA^-^RO^+^CD27^+^ CCR7^+^ (not shown).

**Fig. 2.**
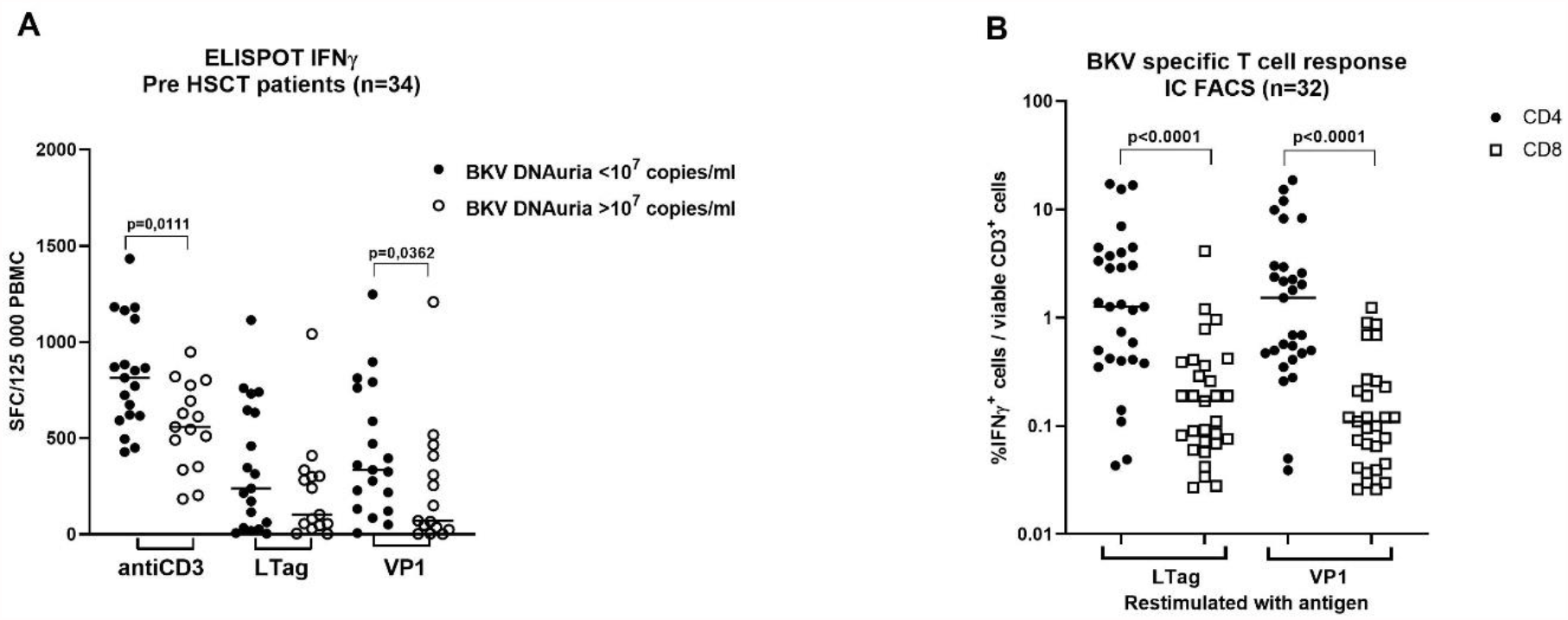
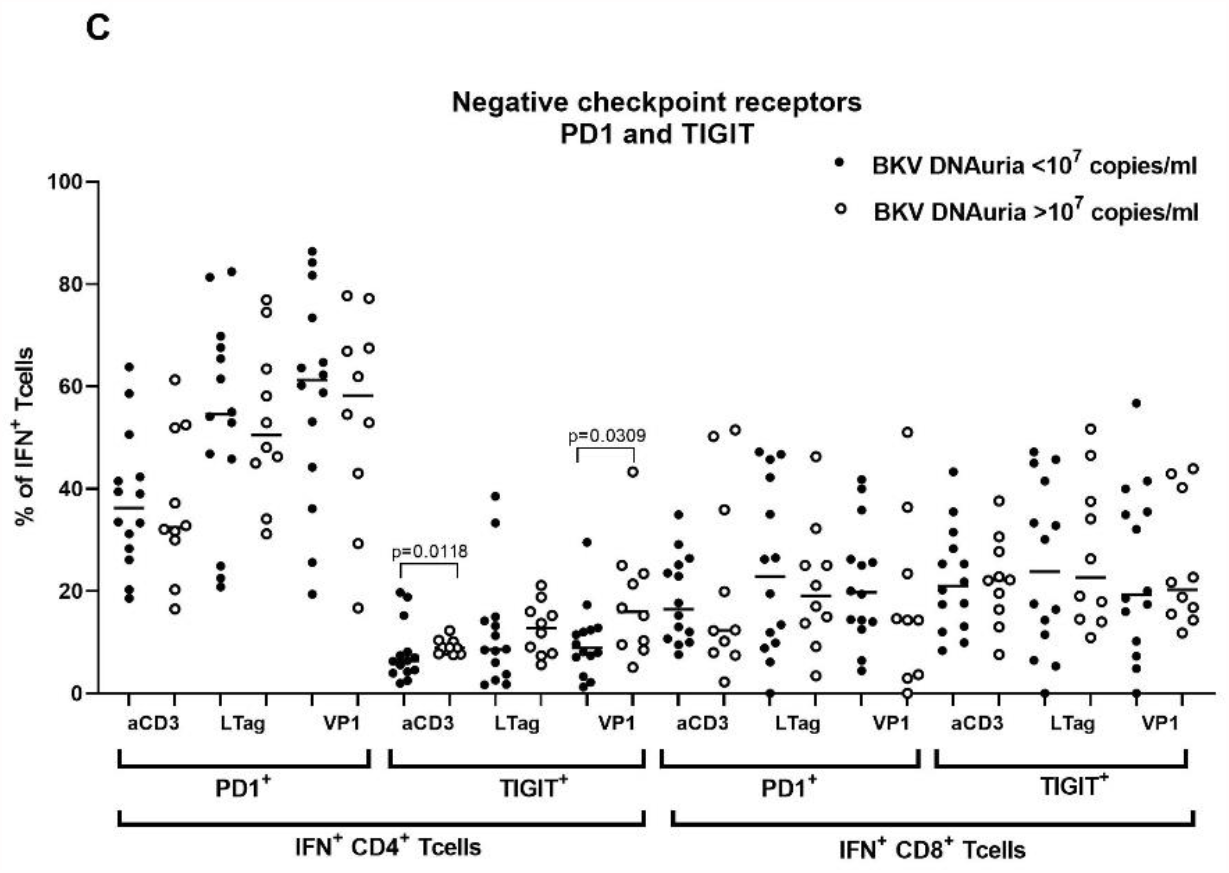
Pretransplant non-specific and BKV specific T cell response in HSCT patients. **(A)** The response to stimulation with anti-CD3 and BKV antigens VP1 and LTag was determined in a subgroup of BKV infected patients by ELISPOT-IFNγ (n=34). Patients were stratified according to maximal BKV DNAuria levels detected during the first year after HSCT. **(B)** The results of ELISPOT-IFNγ were confirmed by the IC FACS-IFNγ in those patients with sufficient yields od expanded T cells (n=32). **(C)** Presence of negative checkpoint receptors PD1 and TIGIT was determined by FACS in IFNγ producing T cells. Statistical analysis was performed using Mann-Whitney test (A, C, D) or the Wilcoxon matched-pairs signed rank test (B).

## DISCUSSION

We analysed pretransplant clinical and immunological conditions which could be used to predict HC following allo-HSCT. Examination of the influence of clinical conditions in a large cohort of HSCT patients has shown that intensity of conditioning regimen was the most crucial pretransplant factor affecting HC. The association of MAC and RIC regimens with HC was very significantly different in our cohort as proved by uni- and multivariate analysis. In accordance with other studies [23] [24-27], patients receiving RIC regimen had a very low risk of HC. The second risk factor associated with increased HC frequency was male gender (Table S1). Therefore, female HSCT recipients were included in LR cohort. The third important factor was associated with prophylaxis of GvHD in recipients of grafts from MMUD, MUD, MMRD and haploidentical donors given either post-transplant cyclophosphamide (ptCy) or ATG. This treatment was associated with a high incidence of HC in patients receiving MAC and had a low and insignificant effect on the incidence of HC if it was given to patients receiving RIC. We compared the incidence of HC in four groups of recipients of grafts from different sources and given ptCy or ATG. Between these individual groups, the risk of HC was not significantly different if they received an identical conditioning regimen. Other studies have also shown that ptCy and ATG in MAC were more frequently associated with HC than RIC containing the same combination [6, 28]. The group of fully matched siblings (MRD) had the lowest incidence of HC due to not so intensive GvHD prophylaxis. A lower HC incidence in the group of MRD in comparison with unrelated donors has been reported by many studies [29-33]. Therefore, male patients in our study receiving any GvHD prophylaxis under MAC regimen were designated as SR. All females, patients receiving RIC regimen and patients with profylaxis lacking ATG or PtCy (MRD) were designated as LR. Incidence of GvHD had no effect on HC occurrence in our cohort.

Our data (Fig. S1B) confirmed the previous findings that BKV viruria correlates with the pre-HSCT anti-BKV IgG level [13] and that BKV viruria >10^7^ copies/ml after HSCT is a very significant risk factor of HC (Table S1 and Table 1) [9, 14]. However, BKV DNA load is not useful as a pretransplant predictor of HC.

The analysis of our data has shown that the risk of HC was higher in younger than in older patients (Table S1). Younger age of adult patients as risk factor for HC was reported by several studies [6, 30, 34, 35]. We believe that predictive power of this association in our study cohort is questionable, given the significantly higher use of MAC than RIC (p<0.0001) in younger patients. Older patients (>51 years) received RIC in 57% of cases whereas younger patients (<51 years) in 18% of cases only. In subgroup of patients who received MAC, the median of age of patients with HC (43 years) was not statistically different from that of patients without HC (45 years) (P=0,0856). This could be explained by age distribution in our cohort as in comparison with other studies the median of age of all patients in this group was much higher.

As a second approach to the prediction of HC, we used the measurement of BKV specific immune response before the start of the conditioning regimen. As for the antibody response, we observed that HC risk was reduced in patients with the lowest anti-BKV IgG levels. This was related to the fact that the group comprised seronegative uninfected patients (OI<1). Patients with anti-BKV IgG levels just above OI value of 1 but below the T1 threshold also had a low incidence of HC. We hypothesised that low levels of anti-BKV IgG in these individuals could mirror a decreased pretransplant tendency of BKV to reactivate and are possibly due to the waning of antibodies. “Medium” anti-BKV IgG levels between T1 and T2 were associated with the highest risk of HC for the group of recipients with significant risk from clinical factors (SR). HC was less common in patients with the highest anti-BKV IgG levels exceeding 75% quartile of positive samples. Predictive value of pretransplant BKV specific IgG was confirmed on the level of virus genotypes. Our results are consistent with a study of humoral responses determined by VLP ELISA (Viracore) in pediatric HSCT patients where the highest pretransplant anti-BKV IgG titers ≥1:163 840 were protective against later BKV viremia and HC whereas IgG titers 1:10 240 were associated with high risk of BKV viremia [15]. On the other hand, Lee et al. [14] working with the same VLP-ELISA reported that increase in BKV IgG titers correlated with developing BKV viruria ≥10^7^ copies/ml and highest BKV IgG titers were not associated with protection. Similarly, Wong et al.[13] using an indirect immunofluorescence assay observed that adult patients with pre-HSCT anti-BKV IgG titer 1:10 had viruria levels up to 10^4^ DNA copies/ml whereas IgG titers >1:20 were connected with viruria ≥10^8^ DNA copies/ml. In their study, very high levels of IgG had no protective effect.

The data on anti-BKV response in donors are rare. Our observation that immune response of donors did not significantly affect BKV reactivation was in line with results of Wong (2007)[13] who reported that donor’s BKV serologic findings did not correlate with BKV viruria in the recipient. From our data it seems that donor/recipient serological mismatch (D-/R+) reported as an important risk factor for cytomegalovirus reactivation in HSCT patients has not a significant effect in case of BKV [36, 37].

To our knowledge, the role of the pretransplant BKV specific T cell response in the protection from BKV reactivation has not been studied so far in HSCT recipients. However, the relevance of posttransplant anti-BKV T cell response for the protection was unequivocally proven by the beneficial effect of adoptive transfer of expanded BKV specific T cells [10] or of donor lymphocyte infusion to patients after HSCT [38]. The importance of BKV-specific T cells for reducing the risk of BKV replication was repeatedly demonstrated in kidney transplant recipients [39-41].

In our study, pretransplant anti-BKV VP1 CD4^+^ T cell IFN^+^ response was associated with low BKV viruria levels and with low frequency of HC after HSCT. On the other hand, weak BKV VP1 CD4^+^ T cell response was associated with high DNAuria after HSCT. CD4^+^ IFNγ^+^ T cells of these patients had increased expression of TIGIT, a known marker of exhaustion and senescence, that is able to modulate T cell function during chronic viral infection [42-45]. It is known that the T cell response can be altered in haemato-oncological patients in consequence of leukaemia itself [46] as well as a result of intensive chemotherapy. In such cases, T cell suppression could contribute to BK virus escape from latency already before transplantation. However, T cell alteration of haemato-oncological patients is not associated with BKV disease before transplantation, and the unlimited virus replication can start soon after HSCT.

Our study has shown that a usage of a combination of clinical and immunological risk factors can help with an early identification of patients who are at risk of developing BKV disease after HSCT. This prediction algorithm could accelerate the preparation of BKV-specific T cells for possible adoptive transfer therapy and improve follow-up of these patients.

## Data Availability

All data and materials obtained during the study are available in my laboratory.

## ACKNOWLEDGEMENT

We thank P. Steinbergerová for a management of patient blood collection and I. Marešová for a sampling of patient sera. We are appreciative to K. Roubalová for critical reading of this manuscript. This work was financially supported by the grant NV-17/31593A of the Ministry of Health of the Czech Republic European regional development fund (ERDF) project AIIHHP CZ.02.1.01/0.0/0.0/16_025/0007428, OP RDE and project for conceptual development of the research organization N° 00023736.

---

**SUPPLEMENTARY TABLE S1.**
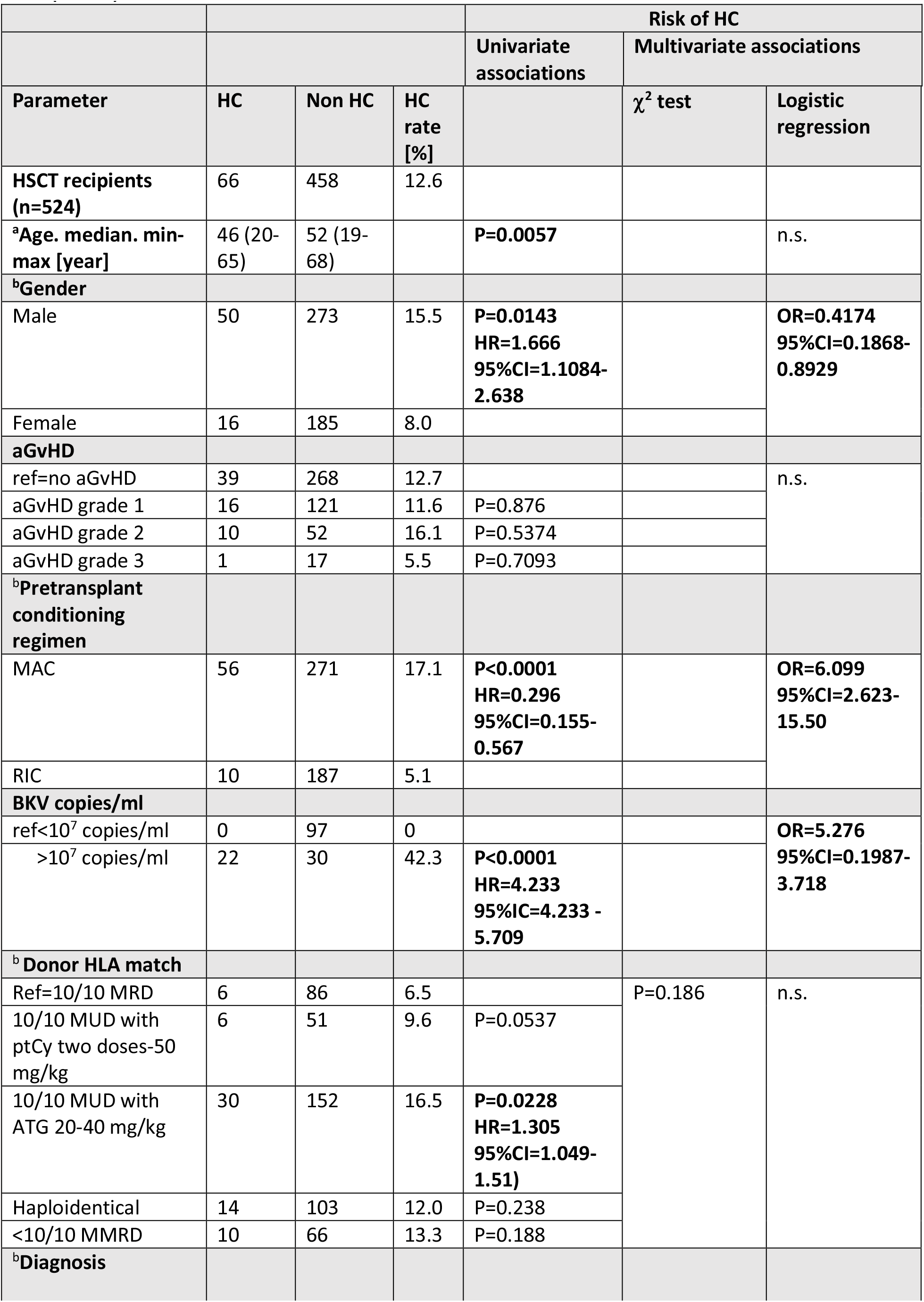
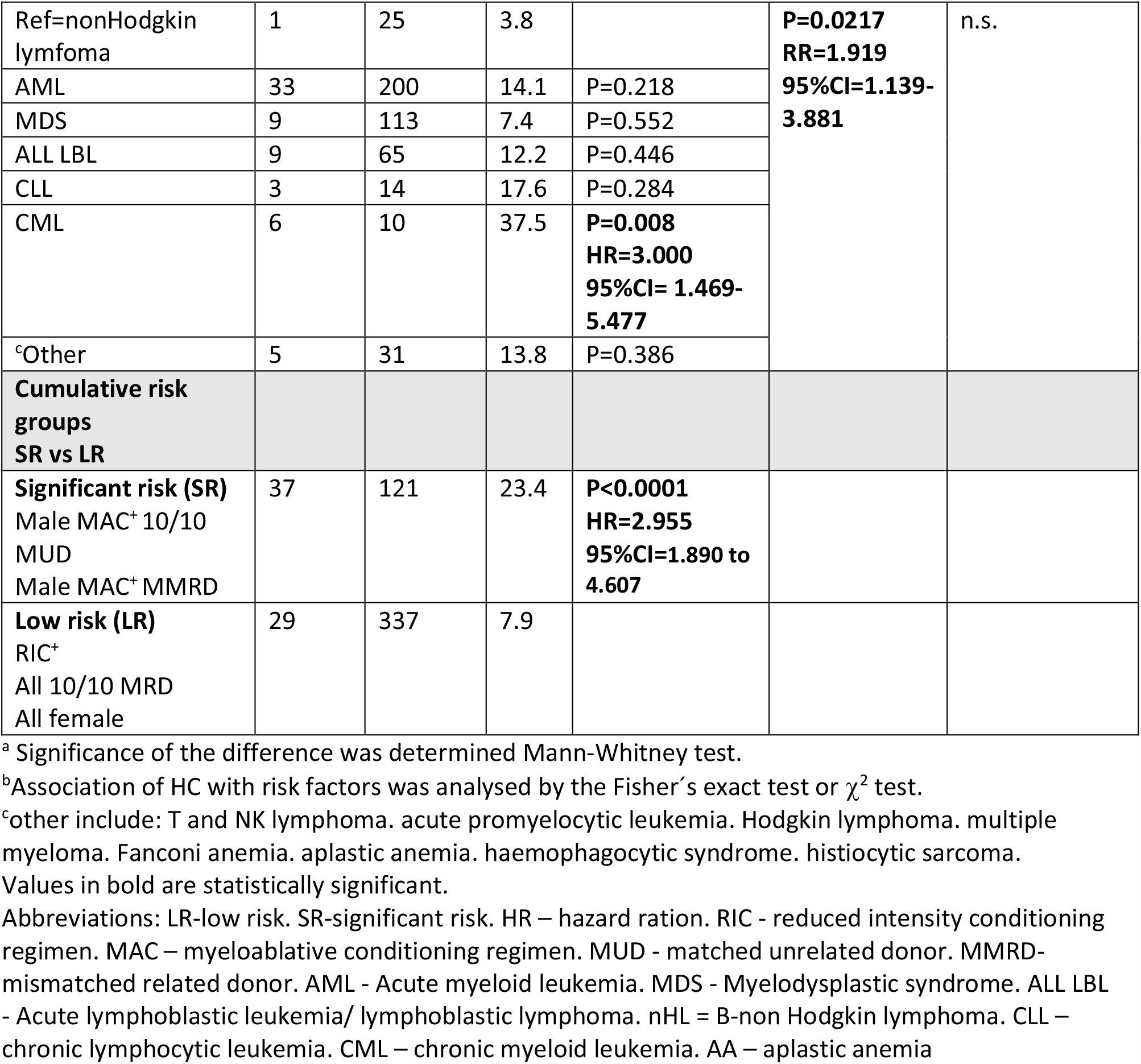
Retrospective analysis of risk of HC in cohort of HSCT recipients transplanted during period 2014 - 2020 (n=524) and clinical features of BKV reactivation.

**Supplementary Figure S1.**
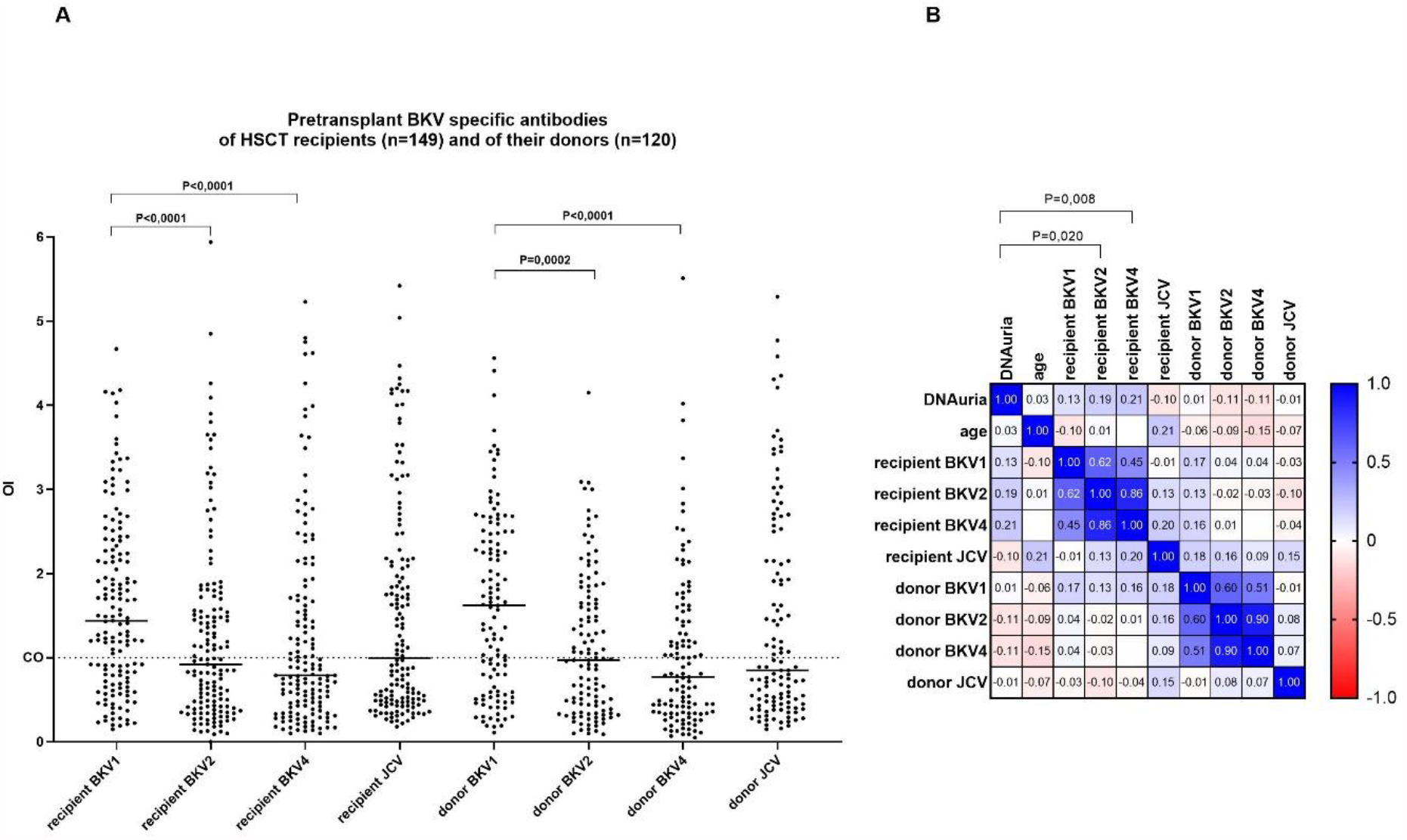
Polyomavirus specific antibodies of HSCT recipients and donors and their relevance for BKV infection. **(A)** Pretransplant antibodies specific for BKV types 1, 2, 4 and JCV were measured in serum or plasma by ELISA. Positive samples have OD index≥1. Significance of differences between median of OD indexes was analysed by paired nonparametric Wilcoxon test. **(B)** Spearman correlation analysis of anti-BKV and JCV IgG of recipients and donors and maximal DNAuria during the first year after transplantation. The heatmap shows Spearman correlation matrix and exact Rs values.

**Supplementary Figure S2.**
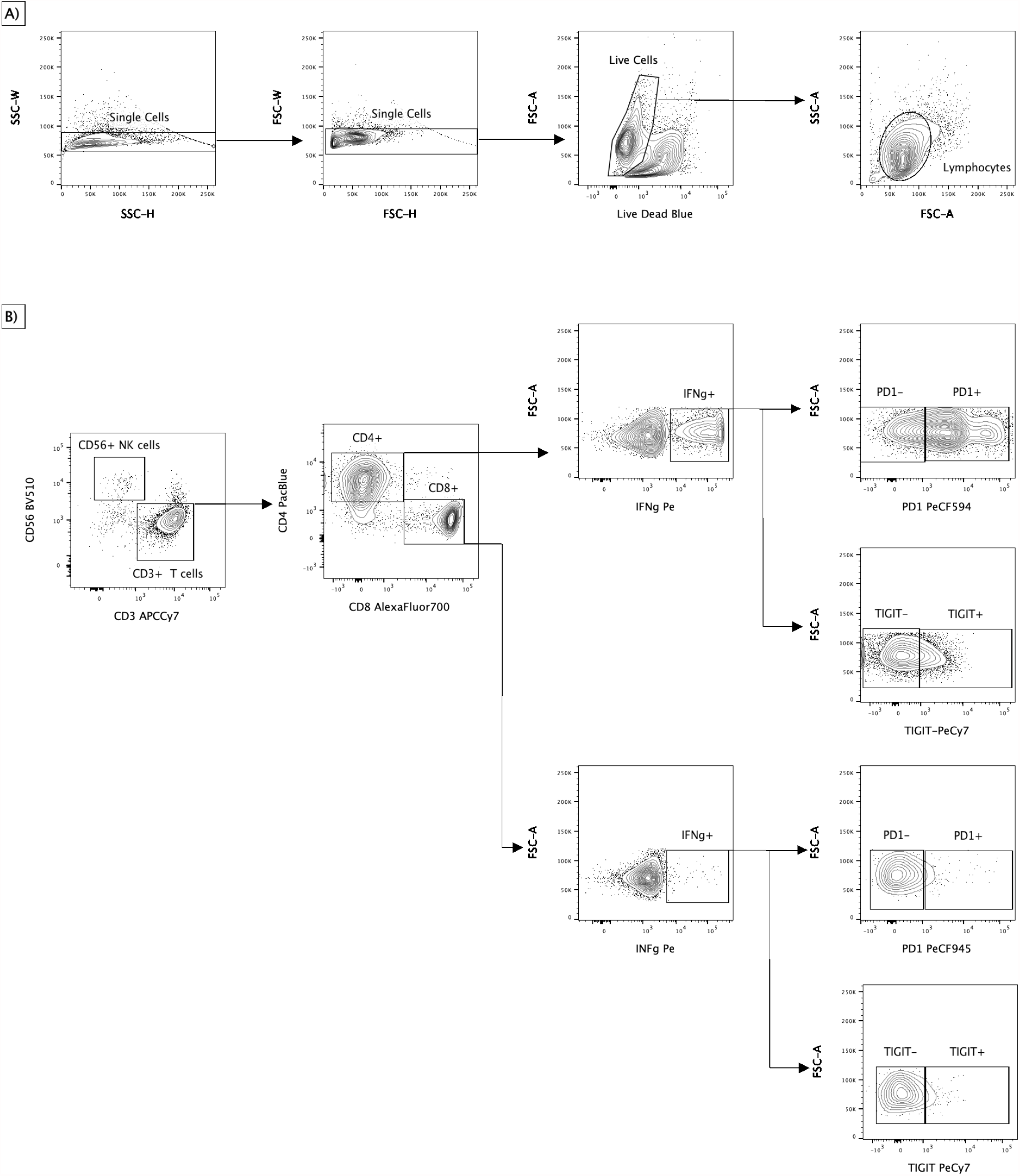
Gating strategy for IC-FACS analysis. **(A)** Doublets, dead cells a debris were removed from the samples and lymphocytes were gated based on SSC and FSC. **(B)** NK cells and T cell were separated using CD56 and CD3. Subsequently expression of IFNγ by CD4 and CD8 T cells was evaluated. Presence of exhaustion markers PD1 and TIGIT was detected on the on CD4+IFNγ+ and CD8+IFNg+ cells.

## SUPLEMENTARY DETAILED DESCRIPTION OF METHODS

### Algorithm for assesment of HC risk upon patient demographic and clinical pretransplant characteristics

The HC incidence in the cohort transplanted between 2014 and 2020 was 13% (Table S1). The values in individual years varied randomly with standard deviation in ±2.92. In univariate analysis the following variables were associated with increased risk of HC: age, male gender, primary diagnosis chronic myeloid leukemia, MA conditioning, MUD transplant with use of ATG. Decreased risk of HC was associated with female gender, reduced intensity (RI) conditioning and MRD transplant. No risk was associated with acute GVHD and prevailing underlying diagnosis such as acute myeloid leukemia (AML), myelodysplastic syndrome (MDS) and lymphatic leukemias. Increased risk of HC was found in patients with chronic myeloid leukemia (CML) (6 of 16) and acute promyelocytic leukemia (2 of 3) which were very rarely treated with HSCT. Based on these pretransplant clinical data patients were divided into low risk group (LR) comprising all female recipients, recipients with RIC and recipients of graft from 10/10 MRD and group with significant risk of HC (SR) comprising remaining patients. Hazard ratio of HC between these two groups differed significantly (Table S1, HR=2.955. 95%CI=1.890 to 4.607. P<0.0001). This method of assessment of the risk of HC based upon clinical characteristics was then used for patients (n=149) in the PBIHC clinical study (Table 1).

#### BKV virus-like particles

The VP1 coding sequences of BKV 1, 2, 4 and JCV, all codon-modified for expression in human cell lines, were synthesized by the GeneArt Gene Synthesis service (Thermo Fischer Scientific). The coding sequences for VP1 proteins of BKV1-D, subtype I/a (, GenBank accession number: JF894228), BKV2-GBR-12 (GenBank accession number: AB263920), BKV4-A-66H, subtype IV/c-2 (GenBank accession number: AB369093), all with a length of 362 amino acids, and JCV with a length of 354 amino acids (strain Mad1. GenBank accession number: J02226.1) were inserted into the pFastBacTM Dual plasmid (Life Technologies) downstream of the polyhedrin promoter. The integrity of the inserted gene was confirmed by sequencing. Recombinant baculoviruses that expressed the VP1 genes were prepared according to the Bac-to-Bac Baculovirus Expression System manual (Life Technologies). Recombinant baculoviruses were plaque purified. BKV VLPs were prepared using the previously described protocols (Sroller et al. 2014) in SF-9 cells. The integrity of the VLPs of individual polyomaviruses was verified by electron microscopy.

### Measurement of BKV specific antibodies

The presence of antibodies to BKV1. 2 and 4 was tested using an in-house enzyme-linked immunosorbent assay (ELISA) based on VP1 virus-like particles (VLP). Briefly, wells of microtiter plates (Polysorp; NUNC, Fisher Scientific, Sweden) were coated with purified BKV1, BKV2 or BKV4 VLPs in PBS (100 ng/well) at 37°C for 2 h and at 4°C overnight. All subsequent incubations were performed at 37°C for 1 h. The wells were repeatedly washed using the automatic microplate washer 1575 (Bio-Rad Laboratories, Hercules, CA) with buffer A (PBS, 0.21 mol/1 NaCl and 0.1% Triton X-100) to remove unbound reagents. Nonspecific binding sites were blocked by incubation with 1% BSA in PBS, and the wells were subsequently incubated in duplicate with human sera diluted 1:100 in buffer A with 1% BSA. Following incubation, antibodies bound were detected with Peroxidase-AffiniPure Donkey Anti-Human IgG (H+L) (Jackson ImmunoResearch Europe Ltd., Ely, UK), and the reaction was visualized by adding 100 μl of o-phenylenediamine containing substrate solution. The colour reaction was stopped by 100 μl of 2 mol/l H_2_SO_4_, and optical densities (ODs) at 492/630 nm were read with the Infinite 200 plate reader (Tecan Trading AG. Switzerland). Background reactivity was determined in wells without antigen. Their absorbances were subtracted from the corresponding values obtained in the presence of the antigen. Positive, negative and cut-off controls for the corresponding antigen were tested on each plate. All ELISA results were represented as a ratio between the absorbance obtained with the tested sample and cut-off control (OD index), which expresses the strength of the antibody response. Samples with an OD index of ≤ 1 were considered nonreactive. All samples within 10% above the cut-off (CO) value, as well as about one-quarter of all serum samples, were retested to confirm the results. Only concordant results 10% above the CO value were classified as reactive. The test was repeated for the third time in the case of discordant results of samples initially reactive/negative. The final result was concordant in two of the three repeated testings.

About 500 sera from healthy blood donors were tested in ELISA with BKV1, BKV2, BKV4 and JCV derived VLPs to distinguish between negative and reactive samples. The CO value was calculated for each antigen as described earlier [4, 21]. Pretransplant antiBKV IgG levels of HSCT recipients were divided into “low”. “medium” and “high” using tresholds T1 and T2 for both BKV1 and BKV4. The threshold T1 was determined as arithmetic mean of seronegative OD index values+3 standard deviations. The value of T1 was 1.35 and 1.25 for BKV1 and 4. respectively. The OD index values lower than T1 comprising non-reactive and “grey zone” samples were assigned as “low” and are plotted in rectangle AZ of Fig 1A. The OD index values laying between thresholds T1 and T2 (plotted in rectangles AY, BY, CY, BX, BZ of Fig 1A were denoted as “medium”. Threshold T2 was determined as 75% quartile value of positive samples (OD>1] and was 2.69 and 2.73 for BKV1 and BKV4, respectively. The values of patients with “high” levels of pretransplant antibodies against BKV1 or BKV4 exceeding T2 threshold were plotted in rectangles AX. CX. CZ in Fig. 1A.

#### Detection of BKV-specific T cells

PBMCs were isolated from the blood of patients and HSC donors on the Ficoll-Paque Plus gradient (GE Healthcare, Uppsala, Sweden). The fresh cells were washed with PBS containing 2% human serum albumin. They were subsequently stimulated for 1 hour with a mixture of 15-amino acid long overlapping peptides (PepMix™) covering BKV VP1 and BKV LTAG (JPT Peptide Technologies GmbH, Berlin, Germany). The PepMixes were used at concentrations of 0.1 µg per 15 million PBMC. The stimulated cells were resuspended in 15 ml of culture medium (CTL) composed of 50% RPMI 1640 with HEPES and glutamine, 45% Click’s medium (both from FUJIFILM Irvine Scientific. Santa Anna. CA), 5% human AB-serum (Capricorn Scientific, Ebsdorfergrund, Germany), 1% Penicillin-Streptomycin-Glutamine (GIBCO, Dublin, Ireland) and supplemented with IL-4 and IL-7 (CellGenics, Freiburg, Germany) at a concentration of 16.6 ng and 10 ng/ml, respectively. They were then cultured in 45 cm^2^ culture flasks (Corning, Corning, NY) under 5% CO_2_ at 37°C. On day 5 of culture, 10 ml of exhausted medium were replaced with the fresh CTL containing 25 ng/ml of IL-4 and 15 ng/ml of IL-7. The culture was completed on the 12^th^ day, and the frequency of the BKV-specific T cells among expanded T cells was measured by ELISPOT-IFNγ or by flow cytometry with intracellular cytokine staining (IC FACS).

#### ELISPOT-IFNγ

Expanded T cells were resuspended in CTL supplemented with 0.5 µg/ml costimulatory molecules (anti-CD28 and anti-CD49d. BD Bioscience) to a concentration of 3.5×10^5^ live cells/ml. From this cell suspension, 0.2 ml/well were seeded in triplicates on ELISPOT plates (MIPN 4550. Sigma-Aldrich, Prague, Czech Republic) precoated with an anti-interferon γ capture antibody (anti-IFN γ, clone 1-D1K, 0.75 µg/well (MabTech, Nacka Strand, Sweden)). Restimulation was performed by individual PepMixes (1 µg/ml) in triplicates under 5% CO_2_ at 37°C for 20 hours. Stimulation with purified anti-human CD3 mAb CD3-2 (anti-CD3) 10 µg/ml (MabTech) was used as the positive control. Cells incubated without stimulants served as the negative control. At the end of the stimulation, the virus-specific T cells (VST) were washed out. IFN γ spots were stained with a biotinylated anti-IFN γ detection antibody 1 µg/ml (clone 7B-6-1-Bi. MabTech), streptavidin-horseradish peroxidase (HRP) conjugate 10 µg/ml (Streptavidin HRP, MabTech) and AEC substrate (BD Bioscience) according to the manufacturer’s protocols. The spot-forming cells (SFC) were counted using the Immunospot analyser (Cellular Technology Limited CTL, Cleveland, OH. USA). The mean SFC numbers were normalized for 1.25 ×10^5^ cells. The SFC values in negative controls were subtracted from those determined in stimulated cultures.

#### Intracellular cytokine detection by flow cytometry (IC FACS)

VST 5 × 10^6^ /ml were stimulated with 1μg/ml PepMix in CTL medium for 2 h and then for an additional 12 h in the presence of 1:1000 diluted Golgi plug (BD Biosciences. Franklin Lakes. NJ. USA). After incubation. cells were harvested with PBS and stained with the LIVE/DEAD Blue Dead Cell Staining Kit (Thermofisher) and antibodies to CD45RA-BB515. CD27-BV650, CD45RO-BV786, PD1-PECF594, CCR7-BV605 (BD Horizon, BD Biosciences), CD56-BV510, TIGIT (VSTM3)-PE/Cy7 (Biolegend, San Diego, CA, USA), CD8-AlexaFluor700 (Exbio, Prague) and CD57 – APC (BD Pharmingen). Cells were then washed with PBS, fixed using IC Fixation & Permeabilization Buffer (eBioscience, San Diego, CA, USA) for 20 min and stained intracellularly with antibodies against IFN-gamma-PE. CD3-APCCy7 (Biolegend) and CD4-PacificBlue (Exbio) in Permeabilization Buffer. Cells were washed and resuspended in FACS buffer (FB - PBS containing 0.09% sodium azide, 1% BSA). Cells were measured using the BD LSR Fortessa 5L flow cytometer (BD Biosciences). The obtained data were analysed by the FlowJo 10.5 software (TreeStar, Ashland, OR, USA). The gating strategy is shown in Fig. S2.

#### BKV genotyping

Viral subtype identification was performed as described by Morel V et al. (2017) [22]. In brief, BKV genotyping was based on a 100-bp segment from nucleotide region 1977 to 2076 within VP1. Viral DNA was first amplified using primers BKV-S1920 5’
s-GGTYATTGGAATAACTAGYATGC-3’ and BKV-A2159 5’-TCCAARTAGGCCTTATGRTCAG-3’. Sanger sequencing of the purified amplicons was performed using the Applied Biosystems BigDye Terminator v3.1 kit according to manufacturer’s instructions. The same primers were used individually for the generation of forward and reverse sequences which were read with the use of Applied Biosystems 3500 Genetic Analyzer. The nucleotide sequences obtained were then manually reviewed and used for subtyping using a described algorithm. DNA of a plasmid pBKV (34-2), subtype BKV1 I/a, kindly provided by J. Forstová, Charles University, Faculty of Science, Prague, served as a positive control.

